# Prodromal Parkinson’s Disease in Essential Tremor: the TITAN study

**DOI:** 10.64898/2026.08.12.26360238

**Authors:** Cristiano Sorrentino, Immacolata Carotenuto, Francesca Di Biasio, Roberto Ceravolo, Matteo Bologna, Nicola Modugno, Salvatore Misceo, Francesca Valentino, Rosa De Micco, Alessandra Nicoletti, Silvia Ramat, Nicola Tambasco, Lazzaro Di Biase, Carlo Colosimo, Anna Rita Bentivoglio, Marinella Turla, Anna De Rosa, Alessandro Stefani, Maria Chiara Malaguti, Carmen Terranova, Francesca Spagnolo, Alessio Di Fonzo, Marcello Esposito, Roberto Tarletti, Laura Brighina, Raffaella Di Giacopo, Mario Coletti Moja, Carlo Dallocchio, Luca Angelini, Enrica Olivola, Angelo Fabio Gigante, Stefan Moraru, Eleonora Del Prete, Laura Avanzino, Andrea Pilotto, Paolo Barone, Roberto Erro, TITAN study group

**Affiliations:** Department of Medicine, Surgery and Dentistry “Scuola Medica Salernitana”, University of Salerno, Salerno, Italy; IRCCS Azienda Ospedaliera Metropolitana, Policlinico San Martino, Genoa, Italy; Department of Clinical and Experimental Medicine, University of Pisa, Pisa, Italy; Department of Human Neurosciences, Sapienza University of Rome, Rome, Italy; IRCCS Neuromed Pozzilli (IS), Italy; Department of Neuroscience and Sensory Organs, Section of Neurology, San Paolo Hospital, Bari, Italy; IRCCS Mondino Foundation Pavia Italy; Department of Advanced Medical and Surgical Sciences, Università della Campania “Luigi Vanvitelli”, Napoli, Italy; Department G.F. Ingrassia, University of Catania, Catania, Italy; Parkinson Unit, Neuromuscular-Skeletal and Sensory Organs Department, AOU Careggi, Florence, Italy; Neurology Department, Perugia General Hospital and University of Perugia, Perugia Italy; Neurology Unit, Campus Bio-Medico University Hospital Foundation, Via Álvaro del Portillo 200, 00128 Rome, Italy; Unit of Neurology, Neurophysiology, Psychiatry, Department of Medicine, Campus Bio-Medico University, Via Álvaro del Portillo 21, 00128 Rome, Italy; Department of Neurology, LUM University, “F. Miulli” Regional Hospital, Acquaviva delle Fonti (Bari), Italy; Movement Disorder Unit, Fondazione Policlinico Universitario A. Gemelli IRCCS, Rome, Italy; Neurology Unit ASST Valcamonica, Esine, Italy; Department of Neurosciences and Reproductive and Odontostomatological Sciences, Federico II University, Naples, Italy; Department of Systems Medicine, University of Rome Tor Vergata, Rome, Italy, Neurology Unit, University Hospital of Rome Tor Vergata, Rome, Italy; Neurology Unit, ASUIT, Santa Maria del Carmine Hospital, Rovereto (Trento), Italy; Department of Clinical and Experimental Medicine, University of Messina, Messina, Italy; Department of Neurology, A. Perrino Hospital, Brindisi, Italy; Neurology Unit, Department of Neuroscience, Dino Ferrari Center, Fondazione IRCCS Ca’ Granda, Ospedale Maggiore Policlinico, Milan, Italy; Clinical Neurophysiology Unit, Cardarelli Hospital, Naples, Italy; SCDU Neurologia - Stroke Unit, Azienda Ospedaliero-Universitaria “Maggiore della Carità”, Novara, Italy; Department of Neurology, San Gerardo Hospital, Monza, Italy; Department of Medicine and Surgery and Milan Center for Neuroscience, University of Milano Bicocca, Milan, Italy; Rovereto Neurology Clinic, University Health Care Service of Trento (ASUIT), Rovereto, Italy; Department of Neurology, Ospedale degli Infermi, Ponderano, Italy; S.C. Neurologia, Dipartimento di Area Medica Specialistica, ASST Pavia, Pavia, Italy; Department of Experimental Medicine, Section of Human Physiology, University of Genoa, Genoa, Italy; Neurology Unit, Department of Medical and Experimental Sciences, University of Brescia, Brescia, Italy; IRCCS Synlab SDN, Naples, Italy

**Keywords:** Essential tremor, Parkinson’s Disease, Prodromal Parkinson’s Disease, Subtle motor signs

## Abstract

**Background:** The relationship between essential tremor (ET) and Parkinson’s disease (PD) remains controversial. Beyond viewing ET as a discrete risk factor for PD, recent frameworks propose that an ET phenotype may represent a clinical presentation of prodromal PD (pPD), consistent with the current reconceptualization of ET as a syndrome. Whether co-occurring subtle motor signs alter pPD probability in ET remains unknown.

**Methods:** Using the MDS research criteria, we calculated pPD probability in a large cohort of ET patients with and without subtle motor signs (rest tremor, hypomimia, isolated rigidity, reduced arm swing, altered repetitive movements, global slowing). Multivariable regression was used to identify independent predictors of pPD probability.

**Results:** Among 599 ET patients (median disease duration: 12 years), only 6 (1.0%) met criteria for probable pPD. Although ET patients with subtle motor signs exhibited higher continuous pPD probability scores than those without, the frequency of possible or probable pPD did not differ significantly between groups. In multivariable regression, neither ET nor individual subtle motor signs, but hypomimia, were independent predictors of pPD probability, which was primarily driven by older age and male sex.

**Conclusions:** Long-standing ET, whether isolated or accompanied by subtle motor signs, is not associated with pPD, with the possible exception of co-occurring hypomimia.

## Introduction

There is longstanding controversy about the relationship between Essential Tremor (ET) and Parkinson’s Disease (PD) [1,2]. Moving beyond the hypothesis of ET as “risk factor” for PD, it has been recently proposed that ET may represent a phenotype of prodromal Parkinson’s disease (pPD) [3,4], a claim that would be consistent with the reconceptualization of ET as a syndrome [5]. However, while a small-scale study of 64 ET patients found that 6.3% met the 2015 criteria for pPD [6], other investigations into individual prodromal features such as hyposmia and rapid eye movement sleep behavior disorder (RBD) yielded variable results [7–11]. This issue is further compounded by the recently proposed ET-plus subtype [5], characterized by the presence of rest tremor and/or of subtle signs. It has been suggested that specific subtle signs might differentially affect the probability of an alternative tremor diagnosis [12–14]. Moreover, subtle motor signs have been reported in the prodromal stage of PD [15–17]. However, whether their presence in ET modulate pPD probability has yet to be thoroughly investigated.

The ITAlian tremor Network (TITAN) is a multicenter data collection platform, aiming to prospectively assess the phenomenology and natural history of tremor syndromes [18]. We here aimed to determine pPD probability according to the Movement Disorder Society (MDS) Research Criteria for pPD [19] within a large ET sample with and without subtle motor signs potentially suggestive of PD, including rest tremor, hypomimia, isolated muscle rigidity, reduced arm swing, altered repetitive movements (e.g., reduced amplitude, velocity or motor halts without the sequence effect) and global slowing.

## Methods

The TITAN protocol has been published elsewhere [18]. For the current study, we identified from the TITAN platform patients with a diagnosis of ET according to the current MDS classification [5] and complete clinical datasets at baseline, including the specific subtle signs contributing to the classification of ET-plus. Patients with subtle signs beyond those listed above (e.g., questionable dystonia, subjective cognitive impairment, impaired tandem gait) were excluded. Risk and prodromal markers for pPD were gathered according to the MDS Research Criteria for pPD [19]. They provide an evidence-based framework for calculating the likelihood of pPD using a naive Bayesian classifier. This approach utilizes age-specific prior probabilities integrated with predictive data from risk and prodromal markers, the latter categorized into motor, non-motor, and biomarkers. The output is expressed as a total estimated likelihood ratio and as a percentage probability of pPD, with a threshold of ≥ 80% required for the diagnosis of “probable pPD” [19]. We also calculated the rate of “possible pPD” using the 50% probability threshold [19] to ensure that the results were not dependent on the latter stricter criteria [20].

Non-parametric tests (e.g., Mann-Withney) and Pearson’s Chi-square test or Fisher’s exact test were performed as appropriate and results are shown as median [interquartile ranges] or percentage distribution. Alpha was set at 0.05 and multiple comparisons corrected with the Benjamin-Hocheberg’s procedure (m =14; Q = 0.05). Finally, a multivariate regression analysis with forward stepping was performed including significant variables in the bivariate analyses and using pPD probability as the dependent variable. All tests were performed using STATA v.18.

## Results

A total of 599 (261 females, 338 males) ET patients, with a median age of 71 [63–77] years, a median AAO of 56 [35–66] years, and a median disease duration of 12 [7–25] years, were extracted from the TITAN database. Among these, 6 patients (1.0%) met the criteria for probable pPD, all of whom were males (6/338 vs 0/261; p=0.032).

Demographic and clinical comparisons between subjects with pET (n=277) and with ET-plus (n=322) are summarized in Table 1. The frequency of subtle motor signs within the ET-plus group was as follow: rest tremor (86.0%); altered repetitive movements (10.6%); rigidity (8.4%); reduced arm swing (3.7%); global slowing (2.5%); and hypomimia (1.6%). At group level, patients with ET-plus were older and showed higher prior pPD probability and prodromal clinical motor and non-motor markers (all p < 0.01; table 1) than patients with pET. While ET-plus patients had a significantly higher total estimated LR, percentage probability and rate of possible pPD than pET, the proportion of probable pPD was comparable between the two groups (Table 1 and supplemental table).

**Table 1.** Clinical comparisons between patients with pure ET and with ET-plus. Abbreviations: AAO: Age-at-onset; LR: likelihood ratio; pPD: prodromal Parkinson’s Disease; TETRAS: The Essential Tremor Rating Assessment Scale. Significant p values are expressed in bold

|  | ET= 277 | ET-plus= 322 | z/Chi<sup>2</sup> value | p |
| --- | --- | --- | --- | --- |
| **Sex (male)** | 58.12% | 54.96% | 0.602 | 0.602 |
| **Age (y)** | 69 [60-76] | 72 [65-77] | -3.75 | **0.000** |
| **AAO (y)** | 54 [30-66] | 57 [43-66] | -1.65 | 0.099 |
| **Disease duration (y)** | 11 [6-26] | 13 [7-24] | -1.17 | 0.239 |
| **TETRAS total score** | 30 [22-40.5] | 35 [25-47.5] | -3.74 | **0.000** |
| **Prior Probability** | .025 [.02-.03] | .03 [.02-.04] | -3.25 | **0.001** |
| **Risk Markers** | .98 [.82-1.26] | .96 [.8-1.29] | -0.23 | 0.821 |
| **Prodromal Markers** | .44 [.2-1.37] | .78 [.28-3.04] | -4.41 | **0.000** |
| **Motor markers** | 1.13 [.48-2.54] | 1.39 [.62-4.4] | -3.02 | **0.003** |
| **Non-motor markers** | .55 [.55-.55] | .55 [.55-.55] | -4.25 | **0.000** |
| **Biomarker** | .66 [.66-.66] | .66 [.66-.66] | -1.20 | 0.229 |
| **LR** | .45 [.22-1.19] | .82 [.31-3.12] | -4.68 | **0.000** |
| **pPD Probability** | 0 [0-3] | 2 [0-9] | -5.67 | **0.000** |
| **Probable pPD** | 0.4% | 1.5% | 2.13 | 0.224 |

Figure 1A illustrates the comparisons between patients categorized by the presence or absence of individual subtle motor signs. Briefly, the presence of rest tremor, hypomimia, rigidity, and altered repetitive movements were associated with an increased pPD probability, that was mostly driven by higher non-motor markers. Indeed, all subtle motor signs but global slowing were associated with higher non-motor markers (figure 1A; supplemental table 1). Conversely, the proportion of probable pPD did not differ significantly (figure 1B) based on the presence of rest tremor (p=0.227), hypomimia (p=0.827), rigidity (p=0.593), reduced arm swing (p=0.725), altered repetitive movements (p=0.546), and global slowing (p=0.775). Similar results were found when applying the 50% probability threshold (supplemental table).

**Figure 1.**
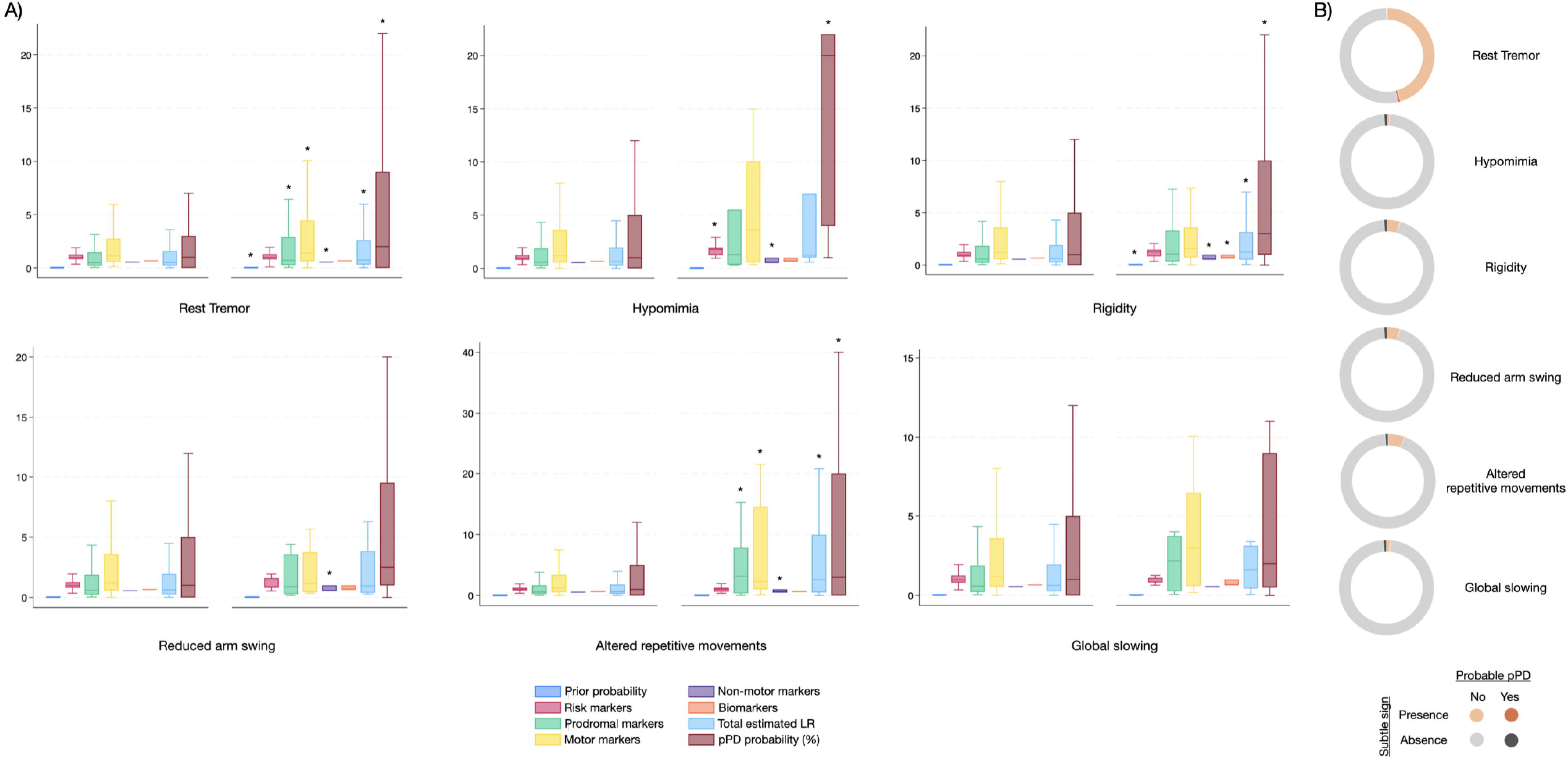
A) Risk markers, prodromal features and pPD probability in patients with (right panels) and without (left panels) individual subtle motor signs. Stars indicate groups with statistically significant higher mean ranks. B) Proportion of probable pPD according to the presence of individual subtle motor signs.

The final regression model (F=15.79; R^2^=0.07; p<0.001), identified age (t=5.69; p<0.01), male sex (t=2.27; p=0.02) and hypomimia (t=2.70; p<0.01) as independent predictors of pPD probability. Other variables, including ET subtype, tremor severity, disease duration, AAO and the presence of other subtle motor signs did not reach the statistical significance.

## Discussion

We sought to calculate the probability of pPD in a large sample of well-characterized ET patients. The prevalence of probable pPD in the entire sample was of 1%, a figure that aligns with the prevalence of pPD in the general elderly population [21]. The comparison between ET subtypes yielded some statistically significant differences, namely higher prodromal motor and non-motor markers and an increased probability of pPD in ET-plus than in pET. Although the frequency of possible pPD was also higher in ET-plus than in pET, the proportion of probable pPD was similar between the two groups and the regression model failed to identify ET subtype as an independent predictor of pPD probability. Similarly, while the presence of individual subtle motor signs disclosed significant differences especially in terms of non-motor markers, which in turn drove a higher probability of pPD, the regression analysis identified hypomimia as the only subtle motor sign independently predicting a higher probability of pPD. Conversely, none of the explored subtle motor signs was associated to a higher proportion of probable pPD.

Prior research investigating the relationship between ET and PD largely focused on individual markers, namely hyposmia or RBD, and yielded variable results, with estimates significantly differing between studies depending on method of ascertainment (self-reported vs laboratory confirmed), cut-offs used for the definition of hyposmia, as well as the criteria for defining ET, which in most instances was based on a retrospective diagnosis [7–11]. The only small-scale study which applied the 2015 criteria for pPD in ET found an increased prevalence of probable pPD (about 6%) that was driven solely by the presence of polysomnography-confirmed RBD as the groups (e.g., with and without RBD) did not differ significantly regarding other prodromal markers [6]. Moreover, this study failed to characterize specific subtle signs and included heterogeneous features like questionable dystonia, cognitive dysfunction, and disequilibrium [6]. Furthermore, their 12% ET-plus prevalence appears underestimated relative to recent literature [22], casting doubts on the finding validity. Our results on a much larger, better characterized and more homogenous ET sample would not support such claims and suggest that the prevalence of probable pPD is in line with the general population of comparable age.

Of note, however, patients with ET-plus as a whole as well as those with specific subtle signs demonstrated an increased pPD probability that was not solely driven by motor markers as one would expect based on the presence of defining features of ET-plus, but mostly by the presence on non-motor markers. Given that ET-plus were also older than ET, it is unclear whether this non-motor profile is specific to the underlying tremor syndrome, to aging or to their combination. Importantly, aging may impact the diagnostic accuracy of pPD criteria, primarily through its interaction with non-motor markers [23]. Future large studies including healthy controls of similar age might answer this question. Similarly, the subtle motor signs that defined the classification of ET-plus (with the exception of rest tremor) might be physiological deviations from normality possibly related to aging, and therefore might represent nuisance features. The only subtle motor sign that was identified as an independent predictor of pPD probability was hypomimia, the low frequency of which might have driven the lack of significant association with probable pPD. Hypomimia is considered an hallmark feature of the disease [24] and, among different parkinsonian signs, it is one of the most reliable predictors of dopaminergic nigrostriatal denervation [25] and of phenoconversion in pPD [26], which might indirectly support our findings.

We acknowledge that, although the diagnosis of ET was based on established clinical criteria [5], the classification of ET-plus based on the presence of subtle motor signs is subjective in the absence of operational criteria for their definition. Nonetheless, this does not detract from the main finding of the current study that ET as a whole is not associated with an increased prevalence of pPD. In addition, we note that such investigations as quantitative smell testing, polysomnography, and neuroimaging that contribute to the calculation of pPD probability as biomarkers, were performed only if clinically recommended, therefore not being available in all patients. Although it has been shown that the accuracy of pPD criteria decreases when based on risk and clinical markers alone [27], our findings were not significantly affected by using the 50% probability threshold for the identification of probable pPD, supporting our interpretation.

In conclusion, our results do not support a significant association between longstanding ET - whether pure or plus - and pPD, with the possible exception of co-occurring hypomimia, a finding that warrants further validation in longitudinal ongoing studies and external cohorts.

## Data Availability

All data produced in the present study are available upon reasonable request to the authors

## Disclosures

### Ethical Compliance Statement

This was work has been approved by the ethic committee of the coordinator center (University of Salerno; study approval n.33_r.p.s.o._02/10/2020) and written informed consent was obtained from the patients prior to their enrolment. We confirm that we have read the Journal’s position on issues involved in ethical publication and affirm that this work is consistent with those guidelines.

### Financial Disclosures for Previous 12 Months

There are no Financial Disclosures for this article.

## Acknowledgments

We are grateful to the “Fondazione Limpe per il Parkinson ONLUS” for the continuing support in the maintenance of the TITAN portal and for coordinating the administrative activities between centers.

## TITAN study group

Maria Russo MD, Neurology Unit, A.O.U. San Giovanni di Dio e Ruggi d’Aragona, Salerno, Italy

Vittorio Gualtieri, Department of Neuroscience, Rehabilitation, Ophtalmology, Genetics and Maternal Child Health, University of Genoa, Genoa, Italy.

Ludovica Cori MD, Department of Clinical and Experimental Medicine, University of Pisa, Pisa, Italy.

Ferdinando Ambrosio MD, Department of Advanced Medical and Surgical Sciences, University of Campania “Luigi Vanvitelli”, Napoli, Italy

Claudio Terravecchia MD, Department G.F. Ingrassia, University of Catania

Alessandra Govoni, MD, Parkinson Unit, Neuroscience and Sensory Organs Department, Azienda Ospedaliero Universitaria Careggi, Firenze.

Pasquale Nigro, MD, PhD, Movement Disorders Center, Neurology Department, Perugia General Hospital and University of Perugia, Perugia, Italy

Pasquale Maria Pecoraro MD, 1 Neurology Unit, Campus Bio-Medico University Hospital Foundation, Via Álvaro del Portillo 200, 00128 Rome, Italy. 2 Unit of Neurology, Neurophysiology, Psychiatry, Department of Medicine, Campus Bio-Medico University, Via Álvaro del Portillo 21, 00128 Rome, Italy.

Marta Filidei MD, Department of Neurology, Santa Maria University Hospital, Terni, Italy

Gabriele Riccio MD, Department of Neurosciences and Reproductive and Odontostomatological Sciences, Federico II University, Naples, Italy

Tommaso Schirinzi MD, PhD, Department of Systems Medicine, University of Rome Tor Vergata, Rome, Italy; Neurology Unit, University Hospital of Rome Tor Vergata, Rome, Italy

Ruggero Bacchin MD, Neurology Unit, ASUIT, Santa Chiara Hospital, Trento, Italy

Antonio Scarola MD, Department of Clinical and Experimental Medicine, University of Messina, Messina, Italy Augusto Rini, Neurological Department, A. Perrino’s Hospital, Brindisi, Italy

Assunta Trinchillo MD, Department of Medical, Motor and Wellness Sciences, University “Parthenope”, Naples, Italy Patrizia Sucapane, MD, Neurology Unit, San Salvatore Hospital, Coppito, AQ, Italy

Susanne Buechner, MD, Provincial Hospital of Bolzano (SABES-ASDAA), Lorenz Boehler Street 5, 39100 Bolzano-Bozen (BZ), Italy.

Sara Varanese MD, Neurology Unit, PO S. Pio da Pietrelcina, Vasto, Italy

Laura Maria Raglione MD, SOC Neurologia Firenze - Azienda USL Toscana Centro Anna Castagna, MD IRCCS Don Gnocchi Foundation, Milan, Italy

## Authors’ Roles

1: conception and design of the study, or acquisition of data, or analysis and interpretation of data; 2: drafting the article or revising it critically for important intellectual content;

3: final approval of the version to be submitted.

- CS: 2,3
- IC: 1,3
- FDB: 1,3
- RC: 1,3
- MB: 1,3
- NM: 1,3
- SM: 1,3
- FV: 1,3
- RDM: 1,3
- AN: 1,3
- SR: 1,3
- NT: 1,3
- LDB: 1,3
- CC: 1,3
- ARB: 1,3
- MT: 1,3
- ADR: 1,3
- AS: 1,3
- MCM: 1,3
- CT: 1,3
- FS: 1,3
- ADF: 1,3
- ME: 1,3
- RT: 1,3
- LB: 1,3
- RDG: 1,3
- MCM: 1,3
- CD: 1,3
- LA: 1,3
- EO: 1,3
- AFG: 1,3
- SM: 1,3
- EDP: 1,3
- LA: 1,3
- AP: 1,3
- PB: 1,3
- RE: 1,2,3

