## Supplemental Table 1 for "Prodromal Parkinson’s Disease in Essential Tremor: the TITAN study"

**Sorrentino C, et al. 2026**

**Supplemental table 1. Distribution of possible prodromal PD (e.g. percent probability > 50%)**

| | Possible pPD YES | Possible pPD NO | p value |
|---|---|---|---|
| **ET/ET+** | 2.16% vs 6.89% | 97.84% vs 93.11% | **0.005** |
| **Rest Tremor (yes/no)** | 4.69% vs 5.17% | 95.31% vs 94.83% | 0.777 |
| **Hypomimia (yes/no)** | 20.0% vs 4.86% | 80.0% vs 95.15% | 0.226 |
| **Rigidity (yes/no)** | 7.41% vs 4.87% | 92.59% vs 95.13 | 0.639 |
| **Reduce arm swing (yes/no)** | 0% vs 5.06% | 100% vs 94.94% | 0.539 |
| **Altered repetitive movements (yes/no)** | 2.94% vs 5.08% | 97.06% vs 94.92 | 0.485 |
| **Global slowing (yes/no)** | 0% vs 5.03% | 100% vs 94.97% | 0.663 |
